# TCIA Radiology Image Processing for AI and Radiomics

**DOI:** 10.64898/2026.06.15.26354651

**Authors:** Joseph Rich, Raphi Kang, David Jin, Saanvi Subramanian, Vinay Duddalwar, Lior Pachter

## Abstract

We developed a standardized, reproducible preprocessing framework for computed tomography (CT) imaging data from multi-institutional repositories such as The Cancer Imaging Archive (TCIA), enabling consistent radiomics and artificial intelligence (AI) analyses. Imaging data from TCGA-KIRC patients available on TCIA were used as a representative heterogeneous dataset characterized by variation in acquisition protocols, inconsistent metadata, and differing image quality. The pipeline includes series filtering, DICOM-to-NIfTI conversion, orientation harmonization to a canonical coordinate system, voxel spacing normalization, intensity clipping and normalization, segmentation integration, and metadata validation, and is implemented in a reproducible, notebook-based framework compatible with common radiomics and deep learning workflows. This pipeline standardizes imaging data into analysis-ready volumes with consistent geometry, intensity distributions, and spatial alignment, reducing non-biological variability that can adversely affect radiomic feature stability and model performance. The modular design enables task-specific adaptation of individual preprocessing steps while maintaining overall consistency. Although demonstrated on TCIA, this framework is generalizable to other heterogeneous imaging datasets and provides a foundation for robust, large-scale computational imaging studies.

## 2 Introduction

Radiogenomics has emerged as an important paradigm in precision oncology, enabled by advances in high-throughput molecular profiling and quantitative medical imaging analysis^1,2^. By linking imaging phenotypes to genomic and transcriptomic alterations, the field aims to noninvasively characterize tumor biology, predict clinical outcomes, and inform treatment selection^3–5^. Recent progress in radiomics and machine learning has further expanded the scope of this approach by enabling the extraction of high-dimensional features from clinical scans and combining them with multi-omic data. However, these methods rely on imaging datasets that are well-annotated and geometrically consistent^6^. Standardized preprocessing for large-scale machine learning workflows is essential to ensure that downstream associations capture biological signal rather than technical variability.

The Cancer Genome Atlas (TCGA) and its imaging counterpart, The Cancer Imaging Archive (TCIA), are among the most widely used public resources for radiogenomics research in oncology^7,8^. TCGA provides comprehensive clinical annotations and genomic data across multiple tumor types, while TCIA hosts the corresponding radiologic imaging studies spanning modalities such as computed tomography (CT), magnetic resonance imaging (MRI), and others. The paired availability of TCGA/TCIA datasets across multiple cancer types has been foundational to develop and benchmark multimodal deep learning models^9–11^.

Despite its value, imaging data from TCIA can be highly heterogeneous. Scans are collected across many institutions, scanners, and time periods, resulting in differences in acquisition parameters such as slice thickness and reconstruction kernels. Individual patient studies often contain multiple series, including localizers, scout images, and various post-contrast acquisitions, many of which have incomplete or inconsistent metadata. DICOM headers also vary in formatting and completeness. Together, these inconsistencies introduce nonbiological variation that can affect radiomic feature stability and model performance if not properly addressed.

In this work, we present a structured protocol to standardize TCIA CT imaging data for radiomics and deep learning applications (Fig. 1). Our workflow consists of key preprocessing steps: series selection, orientation harmonization, voxel spacing normalization, segmentation, size standardization, intensity handling, and metadata validation. Together, these preprocessing steps produce standardized datasets that improve reproducibility and enable consistent comparisons across studies.

**Figure 1:**
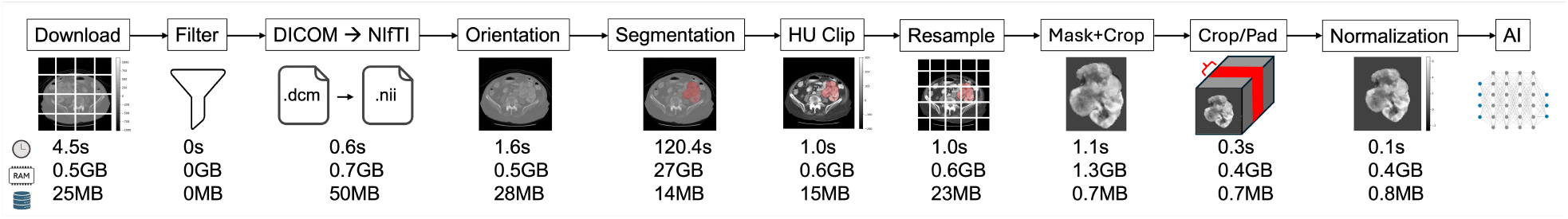
Data processing schematic.

## 3 Methods

We describe the proposed methodology in detail below. The workflow is implemented in a Jupyter notebook with corresponding section titles (see Code Availability), allowing each step to be visualized and debugged interactively.

### 3.1 Data Acquisition

Project-specific metadata were obtained from the TCIA data portal. The required project-level inputs include: (i) `manifest_url`, which contains download links for each imaging series; (ii) `metadata_url`, which provides metadata for all patients, studies, and series; and (iii) `totalsegmentator_organs`, which specifies the organ masks retained from the `--tasktotal` TotalSegmentator output for the cancer of interest.

Metadata were downloaded from the source repository and parsed to summarize key dataset characteristics, including the total number of patients, imaging studies, and individual series. Datasets were categorized by anatomical region and imaging modality, most commonly CT, MRI, or radiography.

Imaging data were downloaded programmatically using the `nbia-data-retriever` commandline utility, which enables reproducible retrieval of DICOM series directly from TCIA using the provided manifest file.

### 3.2 Data Filtering

To generate analysis-ready **axial CT volumes** for radiomic feature extraction and model development, we apply a series of filtering steps to remove non-informative or technically unsuitable imaging series.

#### Modality Restriction

This workflow is designed specifically for computed tomography. Although TCIA archives may contain MRI, PET, or other modalities, only series with `Modality =` “`CT`” were retained. Radiomic features are highly modality-dependent, and voxel intensity interpretation differs fundamentally across imaging types. Restricting the modality ensures consistent Hounsfield Unit–based intensity interpretation, reduces modality-induced variability, and simplifies downstream preprocessing steps such as windowing and resampling. Future work could include providing a standardized protocol for preprocessing non-CT modalities.

#### Exclusion of Series by Metadata Keywords

Series were excluded if any of the DICOM fields `ImageType`, `SeriesDescription`, or `ProtocolName` contained terms associated with non-diagnostic or derived series. The filtering keywords included:

~~~
{“localizer”, “survey”, “asset”, “scout”, “cal”,
“mipseries”, “pjn”, “summary series”,
“topogram”, “mip”, “smart prep”}
~~~

These keywords commonly correspond to localizers, scout and calibration scans, topograms, bolus tracking acquisitions, maximum intensity projections (MIP), or other derived images. Such series typically represent projection images or post-processed outputs rather than native axial diagnostic volumes and are therefore unsuitable for quantitative radiomic analysis.

Series were also excluded if the `SeriesDescription` contained reconstruction kernel identifiers such as “`B5`”, “`B6`”, “`B7`”, “`B8`”, or “`bone`”, which correspond to sharp or high-frequency convolution kernels (e.g., B50–B80). These kernels are optimized for edge enhancement and osseous visualization but substantially increase image noise and high-frequency texture components. Because radiomic features and deep learning models are sensitive to reconstruction kernel differences, mixing sharp and soft-tissue kernels within a cohort can introduce systematic technical variability. Restricting the dataset to standard soft-tissue reconstructions improves feature stability and cross-patient comparability.

#### Slice Thickness Filtering

Series with `SliceThickness > 10 mm` were excluded. Very thick slices are often associated with localizers or summary reconstructions rather than diagnostic axial scans. Radiomic features are sensitive to spatial resolution and voxel anisotropy, and excessive slice thickness increases partial volume effects and reduces textural fidelity. Applying a slice thickness threshold helps preserve meaningful 3D structural information.

#### Minimum Slice Count Threshold

Series containing fewer than five DICOM files were excluded. Extremely small series represent single-slice localizers, partial acquisitions, failed exports, or derived snapshots. In contrast, true diagnostic axial CT volumes generally contain tens to hundreds of slices. A minimum slice-count threshold helps safeguard against incomplete or non-volumetric data.

### 3.3 DICOM to NIfTI

Remaining CT acquisitions were converted from DICOM to NIfTI format to enable standardized volumetric processing and compatibility with common radiomics and deep learning frameworks. NIfTI consolidates an entire imaging volume into a single file while preserving affine geometry and voxel spacing information, simplifying batch processing and spatial transformations. In contrast, DICOM stores each slice as an individual file with distributed metadata. Because most medical image processing libraries (e.g., ITK/SimpleITK^12^, nibabel) and radiomics pipelines natively support NIfTI, conversion facilitates consistent preprocessing and reproducible computational workflows.

#### CT Image Conversion

DICOM series were converted to NIfTI format using `dcm2niix`, which preserves voxel spacing, affine orientation, and metadata while reconstructing the volumetric acquisition into a single 3D file. When multiple reconstructions were present, the axial volumetric acquisition was selected. Axial CT volumes were preferred because they represent the native acquisition plane for most abdominal and thoracic CT protocols and provide consistent geometry for radiomic analysis.

#### Segmentation Conversion and Consolidation

When available, tumor and organ segmentations in DICOM-SEG format were converted to image space using `segimage2itkimage`. Individual label masks were extracted and saved as volumetric NIfTI files. If multiple segmentation objects were present (e.g., left and right organs, tumor and background labels), individual masks were combined into a single multi-label or binary mask volume aligned with the CT image geometry. Consolidating segmentations into a unified mask simplifies downstream processing and ensures spatial consistency with the imaging volume.

#### Removal of 4D Time-Series Data

After conversion, an additional filtering step excluded four-dimensional (4D) NIfTI files. 4D volumes typically represent time-series acquisitions, such as bolus tracking or perfusion monitoring, of a single slice repeated across time points. Because they do not represent full volumetric scans, they are unsuitable for radiomic feature extraction.

#### Exclusion of Non-Axial Volumes via Voxel Spacing Heuristics

To ensure retention of true axial volumetric acquisitions, converted NIfTI files were evaluated using voxel spacing metadata. Volumes with a maximum voxel spacing exceeding 20 mm were excluded. Extremely large spacing along one axis typically indicates coronal or sagittal acquisitions, projection-based series, or partial reformats that erroneously passed earlier metadata filters. In some cases, a series may contain an axial localizer image while the primary volume is acquired in a non-axial plane. Filtering by voxel spacing provides an additional geometry-based safeguard against inclusion of such scans.

### 3.4 Orientation

Orientation refers to the direction in space indicated by each axis (Fig. 2). For instance, the `(R, A, S)` orientation specifies that coordinate values increase toward the patient’s right (R), anterior (A), and superior (S) directions. Each of the three spatial dimensions hasv one of two choices — right/left (R/L), anterior/posterior (A/P), superior/inferior (S/I) — resulting in 8 possible orientations. Anterior refers to the front of the body (toward the chest/face) and posterior to the back, while superior refers to the top of the body (toward the head) and inferior to the bottom (toward the feet).

**Figure 2:**
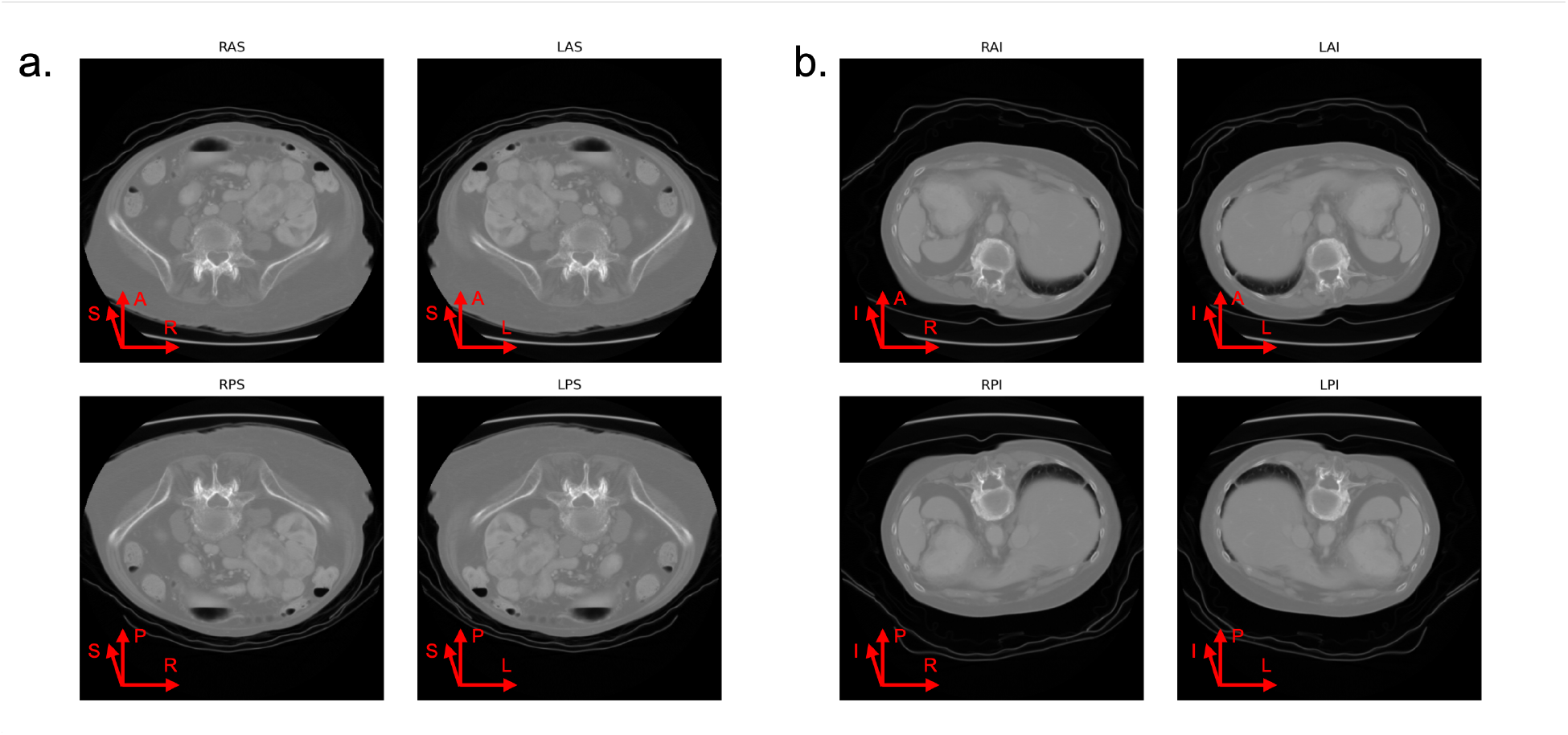
Image orientations. (a) Superior orientations. (b) Inferior orientations.

Following DICOM-to-NIfTI conversion, all volumes were reoriented to a canonical coordinate system using `nib.as_closest_canonical` (NiBabel). Specifically, CT images and masks (when available) were transformed to the `(R, A, S)` convention, corresponding to increasing voxel indices in the Right–Anterior–Superior directions. The `as_closest_canonical` function reorders and flips voxel axes as necessary so that the image conforms to this orientation while preserving spatial correspondence via an updated affine matrix. This operation ensures that voxel axes correspond to the same anatomical directions across all series, enabling consistent spatial interpretation and robust downstream processing.

### 3.5 Organ and tumor segmentation

Segmentation restricts feature extraction to anatomically relevant regions, reducing noise from surrounding tissues that are unrelated to the disease process. For example, in renal cell carcinoma, pathologic changes primarily affect the kidney and tumor itself; including adjacent structures such as the liver can dilute disease-specific imaging signals. Organ segmentation captures morphology and parenchymal characteristics of the affected organ, while tumor segmentation isolates intratumoral heterogeneity and enables region-specific analysis. By focusing analysis on well-defined regions of interest, segmentation improves biological interpretability and reduces variance introduced by irrelevant anatomy.

#### Organ Segmentation

Automated segmentation was performed using `TotalSegmentator`^13^. Small disconnected components were removed using a blob-size filter (`remove_small_blobs`). Organ masks of interest (e.g., left and right kidneys in renal cancer) were selected from the full output set. Post-processing was applied to improve mask robustness and continuity: binary hole filling (`ndi.binary_fill_holes`) eliminated internal voids, and morphological closing (`ndi.binary_closing`) smoothed boundaries and connected adjacent components. From a radiomics perspective, modest over-inclusion is generally preferable to under-segmentation, as exclusion of true organ tissue can bias feature estimates more substantially than mild boundary expansion.

#### Tumor Segmentation

Currently, there is no universally accepted standard for tumor segmentation in TCGA/TCIA datasets. Tumor delineation remains an active area of research, with many task-specific models based on U-Net–style convolutional neural networks^14–17^. Because model performance varies across tumor types, imaging protocols, and annotation standards, tumor masks in this workflow were generated manually under the supervision of an experienced abdominal radiologist to provide high-confidence reference regions. Manual segmentation provides a reproducible baseline for demonstrating the preprocessing pipeline, although automated or semi-automated models may be substituted depending on study requirements.

#### Mask Integration

Organ and tumor masks were combined into a single labeled volume. Voxels corresponding to organ tissue were assigned label value 1 and tumor voxels label value 2, with tumor labels taking precedence in overlapping regions. This unified labeling scheme simplifies downstream radiomic extraction and region-specific feature computation while maintaining spatial alignment with the CT volume.

#### 2D Slice Selection (Optional)

For analyses requiring 2D inputs, a single representative slice can optionally be extracted from the volume. One simple heuristic is to select the slice that contains the largest tumor area, which can be determined using the tumor mask. This approach identifies the slice with the greatest number of tumor-labeled voxels and uses it as the representative image, ensuring that the selected slice captures the most disease-relevant information.

### 3.6 Clipping

CT intensities were clipped to the range [*−* 200, 300] Hounsfield Units (HU) using `sitk.Clamp`. This window approximates a soft-tissue range and captures most abdominal parenchymal and tumor signal while excluding extreme values from air (*< −* 1000 HU) and dense cortical bone (*>* 1000 HU). Clipping reduces the influence of outliers and stabilizes intensity distributions across scans. Alternative clipping ranges may be appropriate depending on the clinical task; for example, lung-focused analyses may use a window near [*−* 1000, 400] HU, while bone-focused studies may require higher upper bounds. The selected range reflects the dominant tissue contrast relevant to the disease under study.

### 3.7 Radiomics

At this point, the images can be passed into a radiomics feature extraction library such as `pyradiomics` if radiomics is an endpoint of analysis. `pyradiomics` is an open-source library implementing standardized quantitative imaging biomarkers^18^. `pyradiomics` automatically performs the subsequent steps of resampling, masking, padding/cropping, and normalization internally. Additionally, radiomic pipelines prefer isotropic spacing (commonly [1, 1, 1] mm), which may differ from the anisotropic spacings native to 3D CT volumes that are generally retained for direct analysis and input into AI models.

#### Feature Extraction Parameters

Several key parameters were specified to promote reproducibility and cross-study comparability. Intensity discretization was performed using a fixed bin width of 25 HU (“`binWidth`”`: 25`). Fixed bin width discretization is commonly recommended for CT radiomics because it preserves the physical meaning of Hounsfield Units and reduces sensitivity to scanner-dependent intensity scaling. Images were resampled internally to isotropic spacing of [1, 1, 1] mm (“`resampledPixelSpacing`”`: [1,1,1]`) prior to feature computation. Isotropic resampling improves rotational invariance and stabilizes texture matrices that depend on voxel neighborhood definitions. B-spline interpolation (“`interpolator`”: “`sitkBSpline`”) was used for intensity images to ensure smooth resampling while minimizing aliasing artifacts. A padding distance of 5 voxels (“`padDistance`”`: 5`) was applied to mitigate edge effects during resampling and filtering operations.

#### Downstream Use

The resulting feature matrix, comprising first-order, shape, and texture-based descriptors, can be used directly for statistical modeling (e.g., survival analysis, regression, or association testing) or as structured inputs to machine learning and deep learning models. Radiomic features provide a compact representation of tumor or organ phenotype that can be integrated with genomic, clinical, or multimodal data in radiogenomic analyses.

All subsequent steps assume the volumes are intended to be passed as input into an AI model.

### 3.8 Resampling

For deep learning workflows, consistent voxel spacing across patients is essential to ensure that convolutional kernels operate over comparable physical distances. Images were resampled to a standardized spacing of (0.8, 0.8, 3.0) mm using `SimpleITK`’s `ResampleImageFilter`.

The output direction and origin were preserved to maintain spatial consistency. Linear interpolation (`sitkLinear`) was used for image volumes, while nearest-neighbor interpolation (`sitkNearestNeighbor`) was applied to segmentation masks to prevent label corruption. Standardizing voxel spacing reduces geometric variability across scanners and acquisition protocols, enabling models to learn anatomy- and pathology-driven patterns rather than resolution artifacts.

### 3.9 Masking

To restrict analysis to the anatomical region of interest, we applied a binary mask derived from the tumor and organ segmentations. Voxels outside the mask were assigned a constant minimum intensity value. The masked image was then cropped to the 3D bounding box of non-minimum voxels with a small padding margin to preserve contextual boundaries while removing irrelevant background.

### 3.10 Cropping/Padding

Deep learning models operating on 3D inputs often require uniform tensor dimensions across samples. To achieve this, target spatial dimensions (*x, y, z*) were defined for the dataset. When appropriate sizes were not known a priori, histograms of the observed image extents were examined and target dimensions were chosen near the 95th percentile of the distribution to capture most volumes while limiting excessive padding.

Each scan was then resized to these standardized dimensions by cropping around the region of interest and applying padding when necessary. This ensures consistent input shapes across the cohort while retaining the relevant anatomical region for downstream modeling.

### 3.11 Normalization

For neural network training, intensity normalization was performed after clipping. Each volume was standardized using

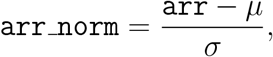

where *µ* and *σ* denote the mean and standard deviation of the image intensities (typically computed per volume). Z-score normalization centers and scales the data, improving optimization stability and preventing gradients from being dominated by intensity scale differences across scans. Unlike radiomics workflows, deep learning models generally benefit from normalized inputs because relative intensity patterns are more important than absolute HU calibration.

## 4 Discussion

The preprocessing steps described provide a practical framework for standardizing multiinstitutional CT imaging data for radiomics and machine learning applications. They address common sources of technical variability in TCGA/TCIA imaging datasets, including heterogeneous metadata, scanner variability, and differences in imaging volume dimensions. However, this pipeline should be viewed as modular rather than universal. Task-specific imaging protocols may require additional filtering steps or alternative preprocessing strategies.

One important example is multiphase contrast-enhanced imaging, which is common in abdominal CT studies such as those involving renal tumors. A single patient may have multiple series corresponding to several phases. Treating each phase as an independent observation can introduce highly correlated scans and artificially inflate sample size. For many radiogenomic applications, it may therefore be appropriate to select a single standardized phase (e.g., venous) to maintain consistency across patients. However, identifying contrast phase is not always straightforward. Phase information may sometimes be encoded in fields such as `SeriesDescription`, but in practice non-contrast series are often absent or mislabeled with inconsistent timing metadata. Under these conditions, unambiguous phase identification may not be possible and may require manual review.

An additional consideration in multiphase imaging is the role of image registration across contrast phases. In renal CT studies, phases such as non-contrast, corticomedullary, and nephrographic are often acquired sequentially, introducing spatial misalignment due to differences in patient positioning, respiratory motion, and temporal changes in organ deformation. When multiple phases are incorporated into a single analysis, registration may be necessary to ensure voxel-wise correspondence across images. Both rigid and deformable registration approaches have been proposed, with deformable methods often better capturing non-linear anatomical changes between phases. However, registration introduces its own sources of variability and potential artifacts, particularly in the presence of large intensity differences between phases. As a result, the decision to apply registration should be guided by the downstream task. For studies explicitly modeling multiphase dynamics, careful registration may be essential, whereas for pipelines that restrict analysis to a single phase, registration can be avoided to reduce preprocessing complexity and potential error propagation ^19^.

Another important consideration in multi-institutional radiomics is the need for feature harmonization to mitigate scanner- and site-specific variability. Even after standardized preprocessing, radiomic features remain highly sensitive to differences in acquisition parameters, reconstruction algorithms, and hardware across institutions, which can introduce non-biological “batch effects” that confound downstream analyses. Statistical harmonization approaches, most notably ComBat and its extensions, have been widely adopted to adjust feature distributions across sites while preserving biologically relevant variation. These methods, originally developed for genomics, have demonstrated effectiveness in reducing inter-scanner variability and improving the reproducibility and generalizability of radiomics models in multicenter settings^20^. However, harmonization is not without limitations: it assumes sufficient representation of each site, may introduce distortions in feature space, and does not fully replace the need for standardized acquisition protocols. Recent work has also explored advanced harmonization strategies, including deep learning–based approaches and covariance-aware extensions, to better capture complex inter-site differences. Public resources such as the CT harmonization multicentric dataset available through TCIA provide valuable benchmarks for evaluating these methods and developing robust pipelines for cross-institutional imaging studies. Overall, harmonization represents a critical complementary step to preprocessing when integrating heterogeneous imaging datasets for radiogenomic and machine learning applications.

In conclusion, we present a reproducible preprocessing framework for preparing CT imaging data for downstream applications. This workflow addresses common sources of heterogeneity and transforms raw TCGA/TCIA imaging archives into standardized, analysis-ready volumes. Although individual studies may require task-specific adaptations, the general principles outlined here could be used for other radiogenomic datasets. As large-scale imaging datasets continue to grow, well-documented preprocessing protocols will remain essential for reproducible and reliable computational imaging research.

## Acknowledgements

J.R. and L.P. were funded, in part, by NIH UM1HG012077.

## Declaration of Competing Interests

V.D. has worked as a consultant for Radmetrix and Roche.

## Data availability

All TCIA imaging data used for this study are publicly available at https://www.cancerimagingarchive.net. The TCGA-KIRC imaging data within TCIA are available at https://www.cancerimagingarchive.net/collection/tcga-kirc/.

## Code availability

All code and Jupyter notebooks can be found on GitHub at https://github.com/pachterlab/tcia-radiology-processing.

